# Evidence for contribution of compound heterozygous variants in Wiskott-Aldrich syndrome like (*WASL*) gene for early onset Parkinson’s disease

**DOI:** 10.1101/2020.08.24.20181024

**Authors:** Sumeet Kumar, Masoom M Abbas, Shyla T. Govindappa, Uday B. Muthane, Sanjay Pandey, B.K. Thelma

## Abstract

**Background:** Knowledge of genetic determinants in Parkinson’s disease is still limited. Familial forms of the disease continue to provide a rich resource to capture the genetic spectrum in disease pathogenesis, and this approach has been exploited in this study.

**Methods:** Informative members from a three-generation family of Indian ethnicity manifesting a likely autosomal recessive mode of inheritance of PD were used for whole exome sequencing. Variant data analysis and *in vitro* functional characterisation of putative disease causal variant(s) identified thereof were carried out in HEK-293 and SH-SY5Y cells using gene constructs of interest.

**Results:** In a rather uncommon observation, two compound heterozygous variants, a rare missense (c.1139C>T:p.P380L) and a novel splice variant (c.1456+5TAGAG>G) in Wiskott-Aldrich syndrome like gene (*WASL*, 7q31), both predicted to be deleterious were shared among the proband and her two affected siblings. *WASL*, a gene hitherto unreported for PD is known to regulate actin polymerisation via Arp2/3 complex. Based on exon trapping assay using pSPL3 vector in HEK-293 cells, the splice variant showed skipping of exon10. Functional characterisation of the missense variant in SH-SY5Y cells demonstrated: i) significant alterations in neurite length and number; ii) decreased ROS tolerance in mutation carrying cells on TBPH induction, and iii) increase in alpha-synuclein protein. Screening for WASL variants in two independent PD cohorts identified four individuals with heterozygous but none with biallelic variants.

**Conclusion:** *WASL*, with demonstrated functional relevance in neurons may be yet another putative disease causal gene for autosomal recessive PD encouraging assessment of its contribution across populations.

## Introduction

A common neurodegenerative disorder generally of the elderly (~1 % in >60 year olds and ~4% in >85 year olds) [1], Parkinson’s disease (PD), is caused by extensive (>50%) loss of substantia nigra pars compacta (SNpc) neuromelanin-pigmented dopaminergic neurons [2]. PD is characterised by motor phenotypes including bradykinesia, tremors and rigidity which are generally preceded by nonmotor symptoms such as depression, anxiety, constipation and sleep disorders [3]. PD is mostly sporadic in occurrence wherein environmental factors along with the genetic components are major contributors, but ~10% of the cases have a Mendelian inheritance pattern suggestive of a stronger genetic underpinning in disease causation. Conventional linkage studies together with contemporary whole exome sequencing strategies using familial PD have identified ~20 genes for PD [4, 5]. Of these, variants in *PRKN*, *PINK1* and *LRRK2* have been most commonly reported in sporadic cases also [6], but with extensive population-specific differences [7–10]. Advances in our current understanding of the disease pathogenesis including proteasome dysfunction, mitochondrial maintenance, vesicular transport and more recently, the involvement of neurodevelopmental and cholinergic pathways; and some insights for newer therapeutic intervention(s) have come from these genetic findings [11]. Conversely, most of the recently identified genes such as *VPS35* and *CHCHD2* are largely private but continue to provide mechanistic insights into PD biology [12, 13]. Mutations in all genes known to date account for <30% of the reported familial cases in most of the populations thus encouraging continued efforts in discovery genomics. To this end, we undertook whole exome sequencing (WES) in a family with three sisters affected with early onset PD. We report for the first time compound heterozygous variants, a rare missense variant (c.1139C>T:p.P380L, MAF=0.000004) and a novel splice variant (c.1456+5TAGAG>G) in Wiskott-Aldrich syndrome like gene (*WASL*, 7q31), as putative disease causal in the study family.

## Methods

### Recruitment of study subjects

A three generation family of south Indian origin (Figure 1) showing a likely autosomal recessive mode of inheritance of PD was recruited from a movement disorder clinic (Parkinson’s and Ageing Research Foundation, India), following the MDS clinical diagnostic criteria [14]. Clearance from the Institutional ethical committee and informed consent from all participants were obtained for the study. In addition, WES data from an independent PD cohort comprising of sporadic young onset (n=205) and familial cases (n=45, with varying age at onset) recruited previously and utilised for other studies were available in the laboratory. Exome data of 120 unrelated healthy individuals and 496 non-PD individuals (mostly schizophrenia cases), all of whom were of Indian ancestry were also available inhouse. WES data from Parkinson’s Progression Markers Initiative (PPMI, www.ppmiinfo.org) PD cohort (n=462) together with 183 healthy controls served as an independent cohort largely of Caucasian ancestry for rare variant screening.

**Figure 1.**
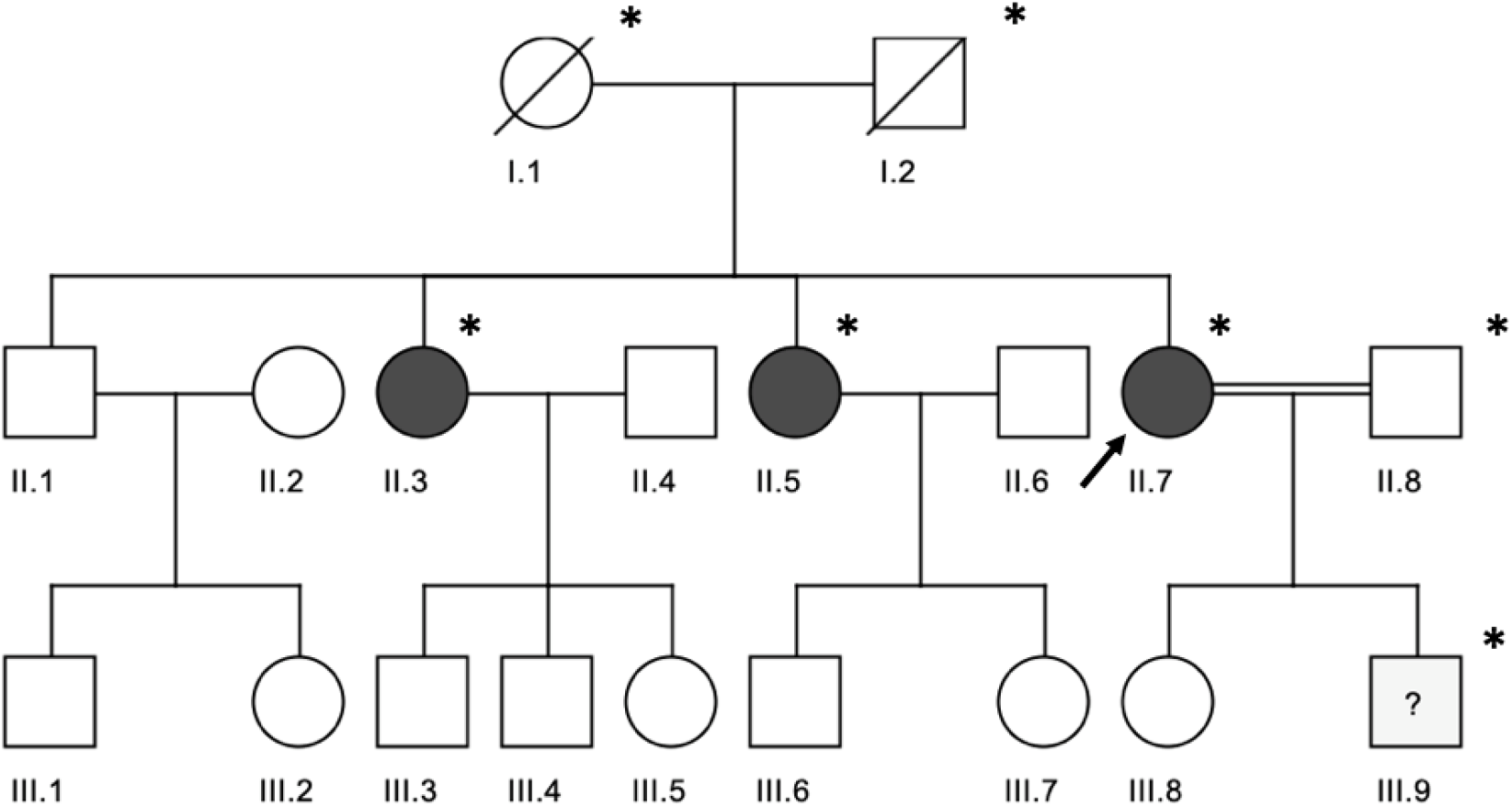
Shows the study pedigree with presumptive autosomal recessive Parkinson’s disease in an Indian family. * whole exome sequenced; arrow indicates the proband;?- III.9 have mild motor symptoms but not yet diagnosed with PD.

### Genetic analysis

DNA isolated from the venous blood of seven family members in the family (Figure 1) was used for exome sequencing.

#### Exome sequencing

Library preparation and enrichment of DNA samples were performed using Agilent V5+UTR kit and sequenced in the paired-end (101 bp) on an Illumina HiSeq2000 at Axeq Technologies, USA (www.axeq.com) and Medgenome Labs ltd, India (www.medgenome.com). Data processing after exome sequencing and variant calling were done using standard bioinformatics pipeline established in the lab as described previously [15]. Variants were annotated using wANNOWAR [16] and prioritised as per standard guidelines [17]. Only rare variants (MAF ≤0.05) shared among three affected individuals were retained. In parallel, kggseq was used for prioritizing the variants [18]. Prioritized variants were then assessed for their segregation profile among all the remaining (not exome sequenced) family members by Sanger sequencing. Exome data were analysed for copy number variations using CNVnator [19].

### *In vitro* functional assays

#### Preparation of construct

Plasmid CB6 with the gene identified based on WES data analysis (detailed in results section) in the family was obtained from Dr. J. Condeelis, Department of Anatomy & Structural Biology, Albert Einstein College of Medicine, New York, USA. Index variant was introduced into the gene construct by site-directed mutagenesis using QuikChangeII kit (Agilent, USA) following the standard protocol (with primers

Fp 5’CCTGGTGGAGGGAGAGGTGGTGGAG3’

Rp 5’CTCCACCACCTCTCCCTCCACCAGG3’).

Plasmid with the gene removed was used as empty vector control (EV).

#### Cell culture

Stable SH-SY5Y cell lines were generated with the wildtype (WT) and variant (VT) constructs of the gene of interest generated as above. Untransfected (UT) SH-SY5Y cells, cells transfected with wildtype ORF (WT) and empty vector (EV) were used as controls.

HEK-293 cells were transfected transiently with cloned wild type and mutant alleles for exon trapping assay (detailed in results section).

#### Western blot

Standard protocols as described previously in the laboratory [20] were followed. Proteins were isolated from the four lines generated as detailed above and resolved on 10% SDS-PAGE. Proteins were detected using Abcam Cat #ab77852 rabbit polyclonal anti-WASL (phospho S484+S485) at 1:2500; anti-alpha synuclein (Abcam Cat #ab52168) at 1:2000; and anti-β-actin at 1:5000) dilutions followed by HRP-conjugated secondary antibody (Abcam) incubation for 1h at room temperature, with β-Actin as the loading control. Quantitation of western blot was performed using ImageJ software.

#### Neurite branching assay

About 8,000 to 10,000 cells from each of the stable SH-SY5Y lines were seeded per chamber on collagen-coated chamber slides (four-well, SPL Lifesciences, Korea). After 24h, the medium was changed to differentiation medium with reduced-serum (1× DMEM+3% serum + 10µM Retinoic acid) and thereafter changed every 48h until the completion of experiment. Cells were differentiated for five days and then stained with a membrane staining dye ‘vibrant dil’ (Thermofisher Cat #V22885). Cells were fixed with 4% paraformaldehyde, prepared for immunofluorescence and images captured at 20X magnification on FLautoIITM fluorescence microscope (Thermofisher, USA). Images were then analysed using IMAGEJ2 software for neurite length per neurite and the total number of neurites per cell. At least 80 cells per category were analysed for both these parameters. All statistical analyses were done using GraphPad Prism trial 7, GraphPad software (La Jolla, California, USA).

#### ROS assay

Cellular reactive oxygen species (ROS) was quantified using the 2’,7’-Dichlorofluorescin diacetate (DCFDA) cellular ROS detection assay kit (Abcam Cat #ab113851) according to the manufacturer’s instructions. Briefly, cells were differentiated as described above. Four hours before ROS measurement, UT, WT and VT lines were treated with 40 µM 6-hydroxydopamine (6-OHDA; Sigma Aldrich Cat #H4381) and then stained with DCFDA for 30 min at 37°C prior to measurements. A similar set of experiments was performed with100 µM *ert*-butyl hydroperoxide (TBPH; Abcam Cat #ab113851). After diffusion into the cell, DCFDA/H2DCFDA is deacetylated by cellular esterases to a non-fluorescent compound, which is later oxidized by ROS into 2’, 7’–dichlorofluorescein which is detected by fluorescence spectroscopy with excitation/emission at 495 nm/529 nm.

## Results

### Clinical details

The proband was a 65year old female symptomatic from the age of 35 years with an insidious onset, progressive motor symptoms of resting hand tremors, slowness, stiffness in the limbs and gait imbalance. On her initial examination (at the age of 50 years, disease duration-15 years), there was no postural hypotension, asymmetrical (left>right) resting tremor, bradykinesia and rigidity in limbs. The pull test was normal. She was managed with a dopamine agonist (piribedil), amantadine and trihexyphenidyl for 4-5 years and levodopa was started later for worsening gait imbalance. She had a good levodopa response, and motor fluctuations were observed over the next 2-3 years. There was a disabling peak dose dyskinesia with dystonia in the great toe and choreiform movements in both feet, even at a low dose (50mg levodopa/12.5mg carbidopa) leading to imbalance and falls. She was reluctant to increase her levodopa dose because of this dyskinesia.

There was a motor progression over the next 15 years, with an increase in her slowness, gait imbalance and freezing. During her latest examination (disease duration-35 years), the extra-ocular movements were full, saccades, speech and swallowing functions were normal. Cognition was normal (Montreal cognitive assessment score-25). There was an asymmetrical limb bradykinesia, rigidity but no tremors. She had gait freezing on turns and the pull test was normal. There were no pyramidal or cerebellar features. The UPDRS III (motor sub-score) one-hour post levodopa (levodopa/carbidopa 50/5) was 35 with lower limb dyskinesia (Supplementary Video 1).

The proband’s two older siblings also have an early onset of Parkinson’s disease (Age at onset: 30 and 41years respectively). The clinical features were similar with a slow motor progression and good levodopa response. There were no autonomic, cognition, or behaviour features; both were independent for their activities of daily living during their last follow up (disease duration-27 and 40 years respectively) (Table 1). Neurological examination of proband’s parents, elder sibling, husband and children was normal.

**Table 1:**
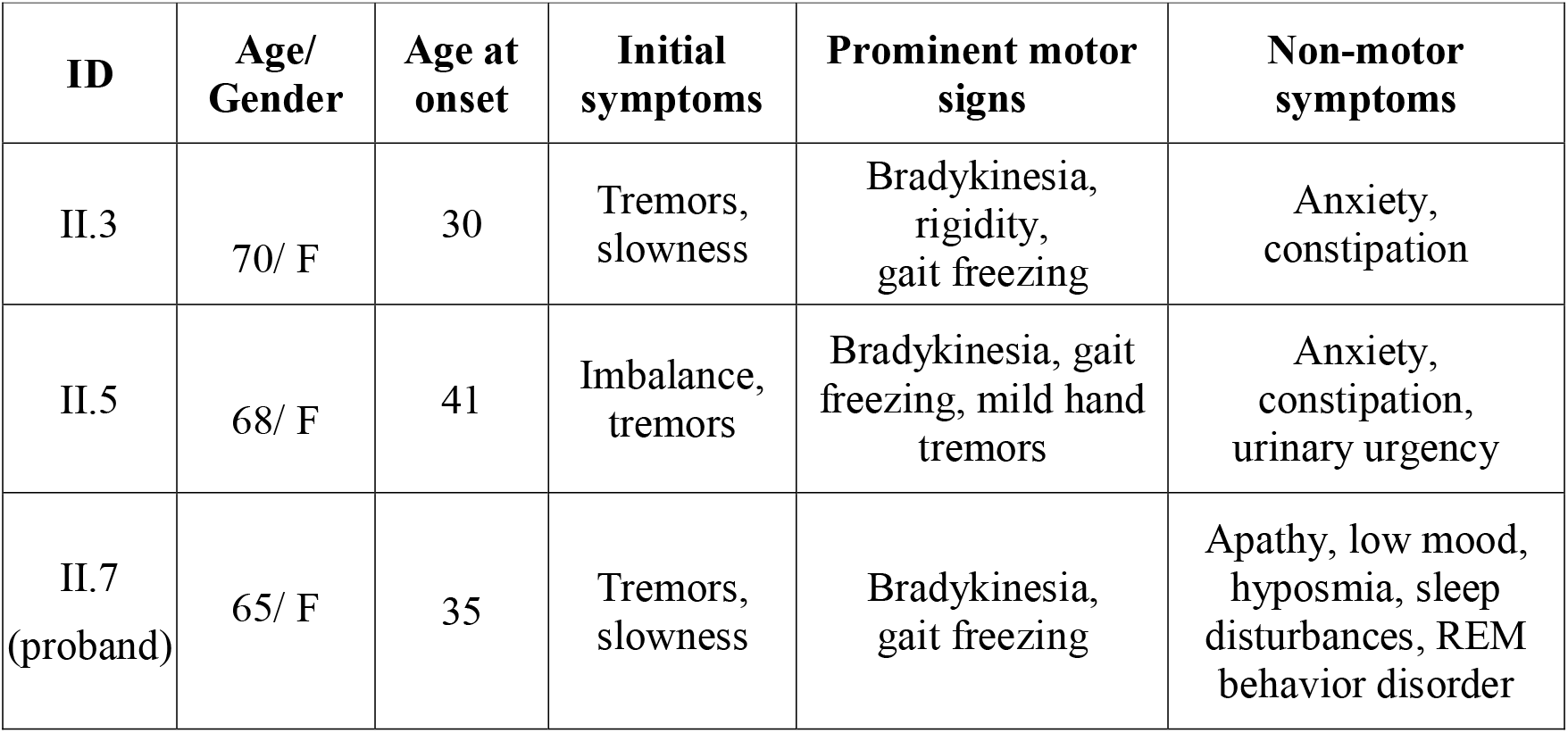
Clinical summary of affected patients

| ID | Age/<br>Gender | Age at<br>onset | Initial<br>symptoms | Prominent motor<br>signs | Non-motor<br>symptoms |
| --- | --- | --- | --- | --- | --- |
| II.3 | 70/ F | 30 | Tremors,<br>slowness | Bradykinesia,<br>rigidity,<br>gait freezing | Anxiety,<br>constipation |
| II.5 | 68/ F | 41 | Imbalance,<br>tremors | Bradykinesia, gait<br>freezing, mild hand<br>tremors | Anxiety,<br>constipation,<br>urinary urgency |
| II.7<br>(proband) | 65/ F | 35 | Tremors,<br>slowness | Bradykinesia,<br>gait freezing | Apathy, low mood,<br>hyposmia, sleep<br>disturbances, REM<br>behavior disorder |

### Genetic Analysis

Copy number variations (CNVs) in *SNCA* and *PRKN* were ruled out in the proband (II.7) using TaqMan and SYBR green assays respectively. Analysis of exome data with CNVnator suggested the absence of CNVs in all other known PD genes as well.

### Whole exome sequencing data analysis

WES of seven individuals (I.1, I.2, II.3, II.5, II.7, II.8 and III.9; Figure 1) yielded a target coverage of 97.8% with >10X depth and 89.3X mean coverage. Variants called therein were annotated using kggseq/wANNOVAR and prioritised, following which, a total of 104669 variants were obtained.

### Compound heterozygous variants in *WASL* segregated in the family

On screening for rare variants shared among the three affected individuals, no homozygous, but 426 heterozygous variants (missense, splice, stopgain and stoploss) were observed. Of these, two heterozygous variants in *WASL* namely, a very rare missense variant c.1139C>T:p.P380L in exon 9 (gnomAD MAF=0.000004) (Supplementary Figure 1A) and a novel splice variant c.1456+5TAGAG>G (exon 10) were found to be shared across the three affected siblings and confirmed by Sanger sequencing (Supplementary Figure 2). Parental screening for these two variants confirmed that the missense variant p.P380L was present only in the father and the splice variant was observed only in the mother. No rare variant(s) in any of the other known PD genes were observed in these siblings. Of note, compound heterozygous variants were also observed in the son (III.9) of the proband, who was born out of consanguineous marriage. Son was not yet diagnosed for PD but had mild motor features like reduced arm swing. Additionally, in third generation DNA samples were available for III.6, III.7 and III.8, screening of which showed the presence of only one variant allele (missense in III.7 and splice variant in III.6 and III.8) and absence of compound heterozygosity.

### Functional characterisation of the index variants

#### *In silico* analysis

Index variants in *WASL* were predicted to be damaging by *in silico* software and the missense variant p.P380L had a CADD score of 22 suggestive of its deleteriousness (Supplementary Table 1). This variant p.P380L also seems to disrupt a highly conserved poly-proline stretch (Supplementary figure 1B), which might lead to structural instability as seen in MD simulation (Supplementary figure 3). Similarly, splice variant c.1456+5TAGAG>G was predicted to cause loss of the donor site by multiple software including Human splicing finder, fruitfly.org and ESEfinder.

#### *In vitro* analysis

The splice variant identified in the family was validated in HEK-293 cells and missense variant p.P380L was assessed for its altered function, if any in SH-SY5Y cells based on neuronal structure parameters.

##### Splice variant

To assess the effect of the 4bp deletion leading to altered splicing using HEK-293 cells, a 1231bp genomic fragment of *WASL* encompassing intron 9, exon 10 and intron 10 from the proband was amplified (primers shown in supplementary table 2) and cloned between two exons of exon trapping vector pSPL3 (Invitrogen, USA) using EcoR1 and XhoI restriction sites. XL1blue cells were transformed with these clones, plasmids isolated from the colonies and screened for wild type and variant alleles by Sanger sequencing (Supplementary Figure 2). A clone each with wild type and variant allele along with an empty vector as control were transfected in HEK-293 cells using lipofectamine 3000. Following transfection, RNA was isolated using TRIZOL reagent and cDNA synthesised using SuperScript VILO cDNA Synthesis Kit (Thermofisher, USA) and used as a template for RT-PCR using primers from pSPL3 exons V1 and V2 (Supplementary Table 2).

##### Variant allele caused exon skipping

RT-PCR products were resolved on a 2% agarose gel in TAE buffer. A 257bp product in the cells with empty vector, a 365 bp product from the wild type allele bearing cells and skipping of exon 10 indicated by a 257bp product in the cells with splice variant were observed (Figure 2).

**Figure 2.**
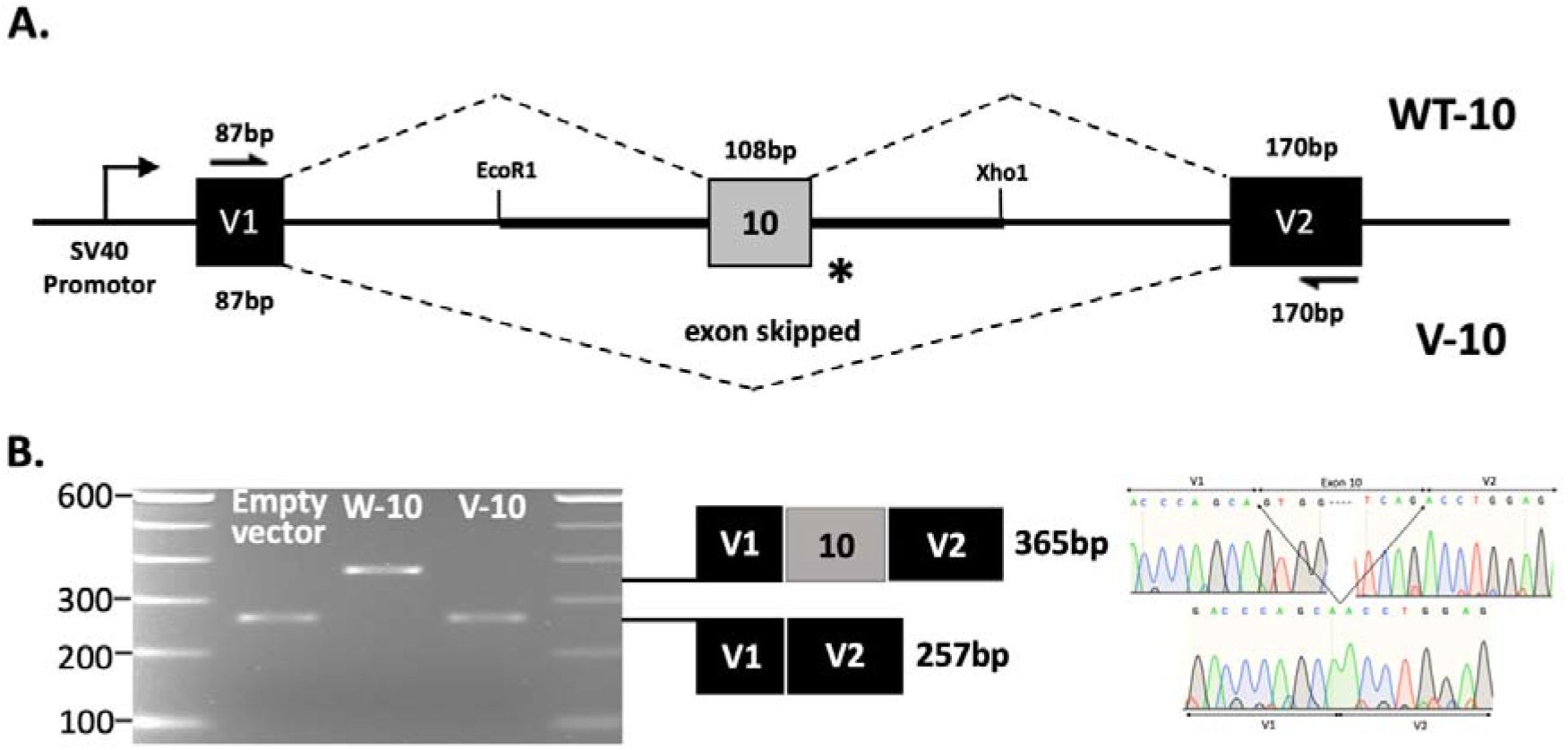
Exon trapping assay. A) A schematic presentation of exon skipping with vector exons V1 and V2 (black boxes) and *WASL* exon 10 (grey). Vector exon-specific primers are indicated by half-arrows. Splicing product of wildtype (WT-10) and variant (V-10) are shown above and below the exon structure (center line) respectively; and B) Gel profile RT-PCR products from transfected HEK293 cells with ‘Empty Vector’ (257bp), WT-10 (365bp) and V-10 (257bp) confirmed by Sanger sequencing as shown by chromatograms.

##### Missense variant

WASL has been shown to be a regulator of actin polymerisation which is indispensable for the structure and function of neuronal networks[21] involved in neuronal structure integrity. Therefore, we used differentiated neuronal SH-SY5Y cells to analyse the role of the p.P380L variant in neuronal structure with neurite length and number as parameters. Additionally, since an excess of ROS is considered to be the common hallmark where multiple pathways converge and lead to cell death in PD, a comparative ROS tolerance of the stable lines generated in the study was assessed following a challenge with ROS inducing compounds. To check if the variant alters alpha-synuclein levels, a well-documented pathway for PD causation, protein levels of alpha-synuclein were also assessed in all the four lines namely, untransfected (UT), wild type *WASL* (WT), variant *WASL* (VT) and empty vector (EV) by western blot analysis.

##### WASL expression confirmed

Protein expression in all the four stable lines was confirmed by western blotting. Two species namely a) ~65 kDa endogenous protein in all the lines and b) ~92 kDa transfected protein (larger size as expected due to GFP tag) only in WT and VT lines but not in UT and EV lines were identified. β-actin was used as a loading control (Figure 3A).

**Figure 3.**
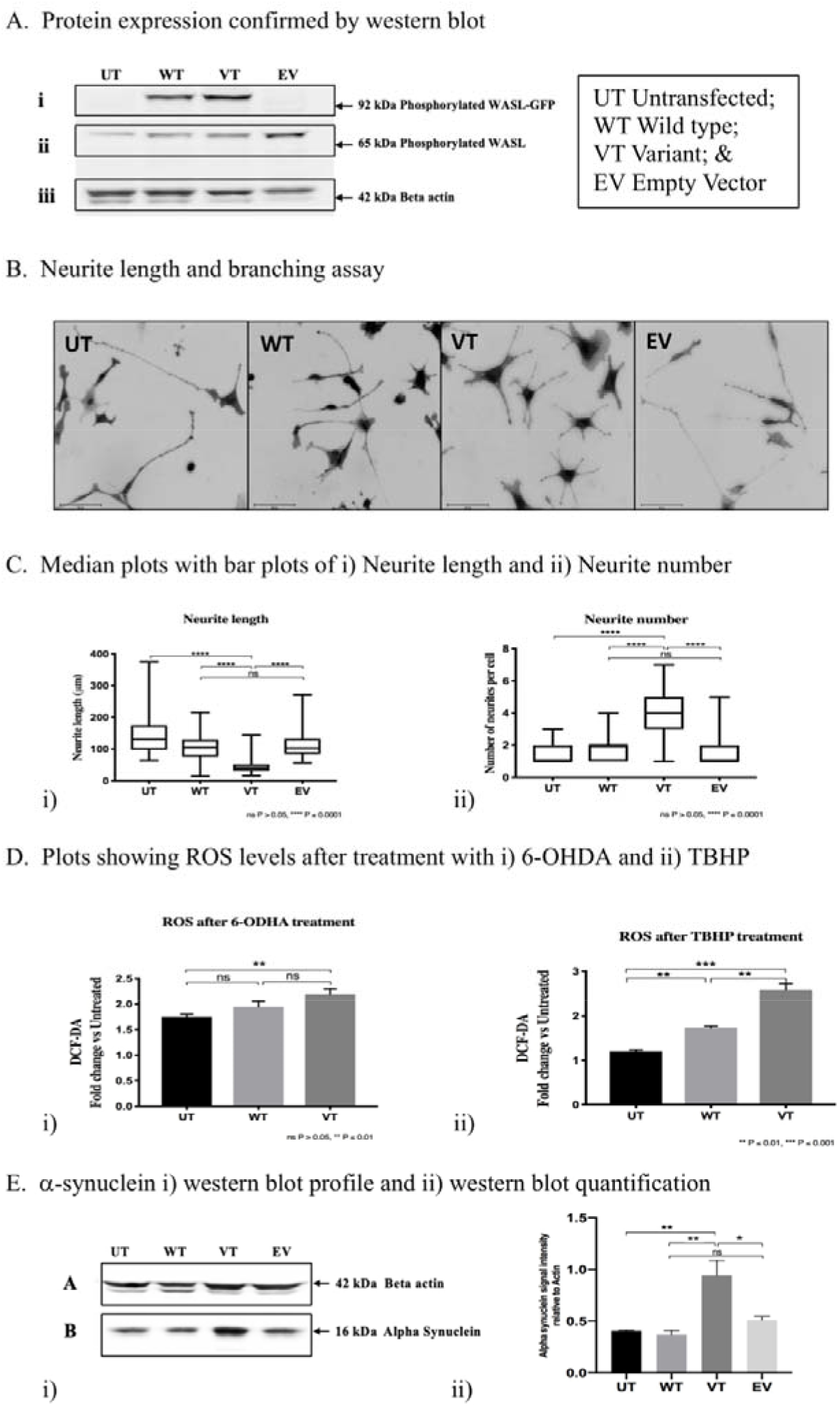
Shows functional assays analysis of *WASL* variant in SH-SHY5Y cells. A, Western blot profile of WASL protein expression using an antibody against phosphorylated WASL showing i) transfected WASL-GFP (~92kDa) only in wild-type (WT) and variant lines (VT); ii) endogenous WASL (~65kDa) in UT, WT, VT and EV lines; and iii) β-actin across all lines. B, Shows differentiated neurons from all four cell lines (UT, WT, VT and EV) stained with membrane staining dye ‘vibrant dil’ V22885 (Thermofisher, USA) to study neurite length and number profiles, with images captured under 20X objective. C, Median plots with bar plots showing significant differences in i) neurite length; and ii) and the number of neurites per cell across the four lines. D, Shows the level of intracellular ROS generated in SH-SY5Y lines UT, WT and VT incubated with a) TBHP (100 μM, 4h) with significant difference in WT and VT line; and b) 6-OHDA (40 μM, 4h) separately and detected by free radical indicator DCFDA (20 μM, 30 min) by fluorescence recorded at Ex/Em of 485 nm and 535 nm, respectively. Data are presented as the ratio of DCF-DA levels in treated vs untreated cells. E, Western blot profile of alpha-synuclein showing a) i) β-actin (~42kDa) used as control and ii) α-synuclein (~16 kDa); b) quantification of western blot where bar plots represent the ratio of alphasynuclein to actin and show a significant difference between VT and other lines.

##### Neurite branching pattern altered in neurons with *WASL* variant

WASL is a brain-enriched regulator of the actin cytoskeleton through Arp 2/3 complex which is involved in the generation of both lamellipodia and filopodia in growth cones and neurogenesis [22]. The functional significance of the missense index variant p.P380L was checked by neurite branching profiling. Total neurite length and the total number of branch points per soma were scored and compared between the UT, WT, VT and EV cells. A significant difference (nonparametric Kruskal-Wallis test with a p-value <0.0001) in both neurite length and number of neurite branch points per cell was observed between VT cells compared to the other three groups (Figure 3B and C i, ii).

##### Neurons with *WASL* variant show less tolerance for ROS

To check if the index variant has any role in ROS homeostasis UT, WT, and VT lines were challenged with two known stress-inducing compounds namely 6-OHDA and TBHP for four hours in independent experiments. Quantity of ROS generated was measured in each line by quantifying the fluorogenic dye 2’, 7’-dichlorodihydrofluorescein di-acetate (H2-DCFDA), which is oxidized by intracellular ROS. After using Tukey’s multiple comparisons, a significant difference (pvalue <0.001) was observed between VT and WT cells with TBHP treatment (Figure 3D ii) but not with 6-OHDA (Figure 3D i).

##### Higher levels of alpha-synuclein was observed in cells with variant

α-synuclein is a key protein where its higher levels have been reported to be involved in PD pathogenesis. To assess its levels, western blot using protein from all four lines was performed. ~16 kDa alphasynuclein band was observed in all four lines along with β-actin (~42 kDa) as a loading control (Figure 3E i). Protein was quantitated using ImageJ software and the ratio of alpha synuclein to β-actin protein were plotted where a significant difference was observed between VT line compared to UT (p=0.008), WT (p=0.006) and EV (p=0.01) lines (Figure 3E ii).

Taken together, these experiments provide preliminary evidence for the functional relevance of both the missense variant c.1139C>T:p.P380L and splice variant c.1456+5TAGAG>G in *WASL* segregating in the study family and thus may contribute to the disease etiology.

#### Screening of independent PD cohorts for additional variants in *WASL*

Screening for variants in *WASL* using available i) in-house exome data for 250 PD patients and ii) publicly available PPMI data for 462 PD cases, identified four different additional rare missense heterozygous variants one in Indian PD case and three in PPMI cases. Of note, in two of the PPMI individuals, a heterozygous missense variant in *LRRK2* and *VPS13C* were also observed (Supplementary Table 1). No homozygous or compound heterozygous variants were observed.

## Discussion

Continued discovery efforts in PD genetics, using vast and distinct patient resources available across different population groups may facilitate confirmation of known and/or enable identification of novel disease causal genes and etiological pathways. Using a hypothesis-free WES strategy, we witnessed compound heterozygous variants in *WASL*, a gene hitherto unreported in PD, shared among three affected siblings in a family of Indian ancestry (Figure 1). The clinical features included an early onset Parkinson’s disease with slow motor progression and a good levodopa response. Interestingly, these variants were also observed in the proband’s son (Aged-35 years), born out of consanguinity (Figure 1). He is asymptomatic and is currently under regular follow up. Identification of four additional rare heterozygous variants on screening two independent PD cohorts, one of Indian origin and the other namely PPMI, largely of Caucasian ancestry (Supplementary Table 1) suggests the likely contribution of *WASL* in PD pathogenesis.

The novel splice variant c.1456+5TAGAG>G observed in the study family predicted to result in the loss of donor site (*in silico*) and skipping of exon 10 was validated in the *in vitro* assays (Figure 2). The second heterozygous rare missense variant c.1139C>T:p.P380L identified in the study seems to affect the highly conserved poly-proline domain (Supplementary figure 1B) which was also functionally characterised by our *in vitro* experiments, demonstrating that this missense change alters the neurite phenotype including decreased length and increased neurite numbers (Figure 3B); and decrease in ROS tolerance in SH-SY5Y cells (Figure 3D). Furthermore, alpha-synuclein protein, whose higher levels have been well established in PD pathogenesis was observed to be expressed highly in *WASL* variant containing cells (Figure 3E). These experimental validations confirm that the compound heterozygous variants identified in the family lead to loss of both functional copies which may be disease causal in the family. Reports of disease causal compound heterozygotes are not uncommon and have been documented in well-established PD genes such as *PRKN* and *PINK1* [23, 24].

WASL via the Arp2/3 complex has been reported to regulate the local rearrangement of actin cytoskeleton thereby inducing the formation of small filopodia, neuronal reshaping of synapses, protrusion of neurites and govern processes involving neural stem cells during embryogenesis and adult neurogenesis [25]. Branching and neurite growth ability in SHSH5Y cells were shown to be affected by the missense index variant in *WASL* (Figure 3B and C) as discussed above. This observation derives support from the reported role of the well-known G2019S mutation in *LRRK2* in shortening the neurite length [26]. WASL has also been previously implicated in endocytosis and myelination of neurons [27, 28]. Many PD associated genes including *SNCA*, *PRKN*, *PINK1*, and *LRRK2* have been implicated in modifying cytoskeleton stability [29]. Role of WASL in triggering actin polymerization during late steps of clathrin-mediated endocytosis and propel clathrin-coated vesicles from the plasma membrane into the cytoplasm has been reported [28]. This vesicular trafficking pathway and its dysregulation has been noted in PD pathology [30]. WASL has also been shown to negatively regulate the transcription of HSP90, which may influence protein folding machinery [31]. In addition to these biochemical studies, *Wasl* knock out mice manifesting PD like symptoms including gait disturbances, dementia [32] and motor deficits [27] lend substantial support to the likely role of *WASL* in PD etiology. Further, the myelinating capacity of cultured rat Schwann cells was impaired by an inhibitor of WASL activity [33]. Benztropine, already in use for treating PD induces remyelination by selectively inducing the differentiation of oligodendrocyte precursor cells [34] linking PD pathology to myelin defects.

Though our *in vitro* experiments only provide evidence for the functional relevance of the variants and not directly to PD pathogenicity, all the studies cited above reiterate the importance of WASL in neuronal function and survival. In summary, compound heterozygous variants comprised of a rare missense and a novel splice alteration in *WASL* segregating with PD phenotype in the study family together with their preliminary *in vitro* functional characterisation provides for the first-time evidence for the likely contribution of *WASL* variants to PD pathogenesis. This study reiterates the importance of continued discovery genomics efforts in transethnic settings to explain missing heritability in PD.

## Data Availability

All data included in manuscript and supplementary deta

## Legends to Supplementary video 1

Patient has gait freezing and lower limb dyskinesia with dystonia of great toes and choreiform movements of feet.

## Acknowledgements

We gratefully acknowledge support from Ms. Anjali Dhyani for maintaining DNA bank in the laboratory; Dr. Jibin John and Mr Surojit Bose, LeadInvent Technologies, India, for assistance in NGS bioinformatics pipeline setup; and Central Instrumentation facility at UDSC for Sanger sequencing services. Infrastructure support provided to the Department of Genetics, University of Delhi South Campus, by the University Grants Commission under the Special Assistance Programme and Department of Science and Technology, New Delhi, under FIST and DU-DST PURSE programs is gratefully acknowledged. We are thankful to Dr. X. Chen and Dr. J. Condeelis, for providing the WASL gene construct. A part of the cohort data used in this manuscript was obtained from the Parkinson’s Progression Markers Initiative (PPMI) database (www.ppmiinfo.org/data) and we gratefully acknowledge them. For up-to-date information on the study, visit www.ppmi-info.org. PPMI, a public-private partnership, is funded by the Michael J. Fox Foundation for Parkinson’s Research and funding partners, including Abbvie, Avid Radiopharmaceuticals, Biogen Idec, Bristol-Meyers Squibb, Covance, GE Healthcare, Genentech, GlaxoSmithKline, Eli Lilly and Company, Lundbeck, Merck, Meso Scale Discovery, Pfizer Inc., Piramal Imaging, Roche CNS group, and UCB.

## Contributors

SK, UBM, MMA and BKT were responsible for the concept and design of the study. MMA, STG, UBM and SP were responsible for diagnosis, clinical characterisation, recruitment and blood collection for all study subjects. SK and BKT acquired, analysed and interpreted the data. SK and BKT wrote the first draft and all authors read the draft, provided their inputs and agreed on the final version of the manuscript before submission.

